# Sociodemographic characteristics and tobacco use patterns associated with using premium, value, and deep-discount cigarettes among US adults who smoke: A cross-sectional analysis of data from the National Survey on Drug Use and Health, 2021

**DOI:** 10.1101/2025.11.18.25340454

**Authors:** Meagan O. Robichaud, Lauren R. Pacek, Michelle T. Bover Manderski, Richard J. O’Connor, Ollie Ganz, Cristine D. Delnevo

## Abstract

Sales of deep-discount cigarettes are growing; yet little is known about who is using these products and their potential impact on tobacco use behavior. Using multinomial logistic regression, this cross-sectional analysis of 2021 National Survey on Drug Use and Health data examines associations between sociodemographic characteristics and smoking behaviors and odds of using value and deep-discount cigarettes (versus premium). Use of value and deep-discount cigarettes was significantly more prevalent for older adults and people with lower incomes and significantly less prevalent among non-Hispanic Black individuals. People who received government assistance, smoked daily, smoked more than a pack of cigarettes per day, and who met the criteria for nicotine dependence were significantly more likely to use deep-discount—but not value—brands than premium cigarettes. More research is needed to better understand how deep-discount cigarettes may impact tobacco use behavior (including smoking cessation), particularly among people with low socioeconomic status.

## INTRODUCTION

Although smoking declined significantly in the United States (US) from 2011-2022, prevalence remained stagnant for those ages 65 and older, and, within this group, it increased among people in the lowest income group.^1^ In recent years, against a backdrop of declining sales, sales of cigarettes in the lowest price tier (i.e., “deep-discount”) have grown.^2,3^ Increasing cigarette prices is a proven strategy to reduce cigarette smoking; however, cheap cigarettes may undermine this.^4,5^ Yet, little is known about who is using deep-discount cigarettes and the potential impact of deep-discount cigarettes on tobacco use behavior. This study examines demographic characteristics and smoking patterns associated with using deep-discount cigarettes.

## METHODS

This cross-sectional analysis of 2021 National Survey on Drug Use and Health^6^ data was considered non–human subjects research by Rutgers University and followed <u>STROBE</u> reporting guidelines. We included adults who smoked cigarettes in the past 30-days and reported a usual cigarette brand (N=6,853), categorizing cigarette brands as: *Premium* if explicitly marketed as “premium” by the parent company, *Value* if offered by major tobacco companies that sell cigarettes in the US (e.g., Altria) and not explicitly marketed as “premium” or “deep-discount,” or *Deep-discount* if a non-premium brand offered by smaller companies or a brand from major tobacco companies explicitly labeled as “deep-discount” (Appendix A).^7,8^ Using Stata version 18 (StataCorp), pairwise chi-square tests assessed differences in demographic and tobacco use variables across brand tiers. We used multinomial logistic regression to examine associations between demographic and tobacco use variables and odds of using value and deep-discount cigarettes (versus premium). Probabilities derived from these odds were used to calculate relative prevalence ratios (RPRs). All analyses accounted for the complex sampling design and were weighted to generate estimates representative of the US civilian, noninstitutionalized population.^9^ We present weighted percentages, RPRs (adjusted for sex, race/ethnicity, income, receiving government assistance, daily smoking, mean cigarettes smoked per day, and nicotine dependence), and 95% confidence intervals.

## RESULTS

Premium cigarettes were most prevalent (77.2%, n=5,607), followed by value (13.2%, n=743) and deep-discount (9.6%, n=503; Table 1). Distributions of all variables (except education) differed significantly across tiers, with value and deep-discount tiers having higher proportions of people who were female, non-Hispanic White, aged 50 or older, had household incomes below $50,000, and received government assistance. Daily smoking, nicotine dependence, and smoking more than a pack per day were most prevalent among individuals using deep-discount cigarettes.

**TABLE 1:** Demographic and Tobacco Use Characteristics of US Adults (2021) Using Premium, Value, and Deep-Discount Cigarettes (n, weighted column %, 95% CI)

|  | **Total Sample**<br>N=6,853 | **Premium**<br>n=5,607, 77.2 [75.1-79.3] | **Value**<br>n=743, 13.2 [11.7-14.9] | **Deep Discount**<br>n=503, 9.6 [8.1-11.3] |
| --- | --- | --- | --- | --- |
|  | **N, % [95% CI]** | **n, % [95% CI]** | **n, % [95% CI]** | **n, % [95% CI]** |
| **Sex** |  |  |  |  |
| Male | 3,326, 52.3 [50.2, 54.4] | 2,772, 54.5 [51.9, 57.1]<sup>A</sup> | 342, 41.8 [35.1, 48.9]<sup>B</sup> | 212, 48.8 [41.3, 56.3]<sup>A,B</sup> |
| Female | 3,527, 47.7 [45.6, 49.8] | 2,835, 45.5 [42.9, 48.1] | 401, 58.2 [51.2, 64.9] | 291, 51.3 [43.7, 58.8] |
| **Race/Ethnicity** |  |  |  |  |
| NH White | 4,415, 67.2 [64.3, 69.9] | 3,464, 62.9 [59.5, 66.1]<sup>A</sup> | 539, 78.5 [72.3, 83.6]<sup>B</sup> | 412, 86.2 [80.3, 90.5]<sup>B</sup> |
| NH Black | 894, 13.5 [12.1, 15.1] | 814, 15.8 [14.1, 17.7] | 65, 7.6 [5.2, 10.9] | 15, 2.9 [1.1, 7.7] |
| Hispanic | 840, 12.5 [10.6, 14.6] | 738, 13.9 [11.6, 16.6] | 69, 8.5 [5.3, 13.3] | 33, 6.2 [3.3, 11.4] |
| NH Other Race | 704, 6.9 [5.6, 8.2] | 591, 7.4 [6.2, 8.9] | 70, 5.5 [3.3, 8.9] | 43, 4.7 [2.6, 8.3] |
| **Age Group** |  |  |  |  |
| 18-25 | 1,628, 8.9 [8.0, 10.0] | 1,439, 10.4 [9.4, 11.5]<sup>A</sup> | 114, 4.4 [2.8, 6.8]<sup>B</sup> | 75, 3.6 [2.4, 5.3]<sup>B</sup> |
| 26-34 | 1,694, 18.9 [17.4, 20.5] | 1,457, 21.4 [19.5, 23.4] | 140, 11.3 [8.2, 15.2] | 97, 9.4 [6.6, 13.1] |
| 35-49 | 2,256, 29.6 [27.5, 31.8] | 1,836, 30.9 [28.3, 33.5] | 255, 26.4 [21.7, 31.8] | 165, 24.1 [17.6, 32.0] |
| 50-64 | 867, 29.5 [26.5, 32.7] | 628, 27.2 [24.1, 30.6] | 140, 36.1 [28.5, 44.5] | 99, 38.8 [29.9, 48.5] |
| 65+ | 408, 13.0 [11.3, 14.9] | 247, 10.1 [8.4, 12.2] | 94, 21.9 [15.8, 29.5] | 67, 24.2 [17.0, 33.1] |
| **Income** |  |  |  |  |
| <\$20,000 | 2,096, 29.5 [27.3, 31.9] | 1,645, 27.4 [24.6, 30.3]<sup>A</sup> | 254, 35.8 [29.4, 42.8]<sup>B</sup> | 197, 38.2 [30.3, 46.9]<sup>B</sup> |
| \$20,000 - \$49,999 | 2,363, 35.5 [32.9, 38.2] | 1,894, 34.4 [31.5, 37.4] | 270, 38.9 [32.2, 46.1] | 199, 39.8 [32.0, 48.2] |
| \$50,000 - \$74,999 | 955, 14.3 [13.0, 15.7] | 797, 15.5 [13.8, 17.2] | 103, 10.0 [7.6, 13.0] | 55, 11.1 [6.4, 18.5] |
| ≥\$75,000 or More | 1,439, 20.7 [18.7, 22.8] | 1,271, 22.8 [20.5, 25.3] | 116, 15.3 [10.9, 21.1] | 52, 10.9 [7.4, 15.7] |
| **Educational Attainment** |  |  |  |  |
| Less than High School | 1,190, 17.7 [15.9, 19.6] | 945, 17.7 [15.5, 20.2]<sup>A</sup> | 142, 16.2 [11.4, 22.6]<sup>A</sup> | 103, 19.2 [13.9, 26.0]<sup>A</sup> |
| High School Graduate | 2,420, 38.1 [35.9, 40.4] | 1,909, 36.9 [34.5, 39.5] | 306, 41.9 [34.7, 49.3] | 205, 42.7 [34.7, 51.0] |
| Some College | 2,255, 32.0 [29.8, 34.2] | 1,869, 32.2 [29.4, 35.2] | 228, 33.3 [26.7, 40.8] | 158, 28.3 [22.4, 34.9] |
| College Graduate | 988, 12.2 [10.5, 14.3] | 884, 13.2 [11.4, 15.1] | 67, 8.6 [4.5, 15.7] | 37, 9.8 [5.3, 17.4] |
| **Receives Govt Assistance** |  |  |  |  |
| No | 4,348, 61.8 [59.8, 63.7] | 3,684, 63.6 [61.4, 65.8]<sup>A</sup> | 419, 58.8 [51.5, 65.7]<sup>A,B</sup> | 245, 50.9 [42.6, 59.2]<sup>B</sup> |
| Yes | 2,505, 38.2 [36.3, 40.2] | 1,923, 36.4 [34.2, 38.6] | 324, 41.2 [34.3, 48.5] | 258, 49.1 [40.8, 57.4] |
| **Days Smoked (Past 30 Days)** |  |  |  |  |
| Non-Daily (<30 Days) | 2,935, 38.3 [36.2, 40.4] | 2,605, 43.1 [40.6, 45.6]<sup>A</sup> | 240, 25.3 [20.4, 31.0]<sup>B</sup> | 90, 17.0 [12.0, 23.5]<sup>B</sup> |
| Daily/All 30 Days | 3,888, 61.7 [59.6, 63.8] | 2,977, 56.9 [54.4, 59.4] | 502, 74.7 [69.0, 79.6] | 409, 83.0 [76.5, 88.0] |

**TABLE 1 (Continued):** Demographic and Tobacco Use Characteristics of US Adults (2021) Using Premium, Value, and Deep-Discount Cigarettes (n, weighted column %, 95% CI)
|  | **Total Sample**<br>N=6,853 | **Premium**<br>n=5,607, 77.2 [75.1-79.3] | **Value**<br>n=743, 13.2 [11.7-14.9] | **Deep Discount**<br>n=503, 9.6 [8.1-11.3] |
| --- | --- | --- | --- | --- |
|  | **N, % [95% CI]** | **n, % [95% CI]** | **n, % [95% CI]** | **n, % [95% CI]** |
| **Mean Cigarettes/Day** |  |  |  |  |
| <10 | 3,726, 48.4 [46.1, 50.7] | 3,286, 54.3 [51.9, 56.7]<sup>A</sup> | 301, 32.4 [25.7, 39.8]<sup>B</sup> | 139, 22.8 [16.5, 30.8]<sup>B</sup> |
| 10 to 19 | 1,678, 25.6 [23.6, 27.6] | 1,274, 23.0 [20.8, 25.4] | 238, 33.2 [27.0, 40.0] | 166, 35.4 [29.0, 42.3] |
| 20 CPD (1 pack) | 1,108, 19.6 [17.4, 22.1] | 814, 17.5 [15.1, 20.3] | 147, 26.0 [19.9, 33.3] | 147, 28.1 [21.4, 35.9] |
| >20 (>1 pack) | 333, 6.4 [5.3, 7.7] | 228, 5.2 [4.1, 6.5] | 54, 8.4 [4.9, 14.2] | 51, 13.7 [8.6, 21.0] |
| **Nicotine Dependent** |  |  |  |  |
| No | 3,315, 45.3 [42.4, 48.2] | 2,902, 49.6 [46.4, 52.9]<sup>A</sup> | 276, 36.9 [31.1, 43.1]<sup>B</sup> | 137, 22.3 [17.3, 28.3]<sup>C</sup> |
| Yes | 3,272, 54.7 [51.8, 57.6] | 2,476, 50.4 [47.1, 53.6] | 443, 63.1 [56.9, 68.9] | 353, 77.7 [71.7, 82.7] |

A, B, C: Categories that do not share superscripts (A, B, or C) refer to statistically significant differences between brand tiers at a Bonferroni corrected alpha of 0.017.

Use of value and deep-discount (versus premium) brands was significantly more prevalent for people 50 and older (versus 18-25) and earning less than $50,000 annually (versus ≥$75,000, Table 2). Use of both brand tiers (versus premium) was significantly less prevalent among non-Hispanic Black than non-Hispanic White people. Females (versus males) and people ages 35-49 (versus 18-25) were significantly more likely to use value than premium brands; however, neither sex nor the 35-49 age group were associated with relative prevalence of deep-discount brands. Individuals smoking 10-19 (versus <10) cigarettes per day were significantly more likely to use value than premium brands, while individuals smoking more than a pack per day were significantly more likely to use deep-discount brands. People receiving government assistance, smoking daily, and with nicotine dependence were significantly more likely to use deep-discount—but not value—brands than premium cigarettes.

**TABLE 2:** Adjusted^†^ Relative Prevalence Ratios (RPRs) and 95% Confidence Intervals (CIs) for Proportion of Individuals Using Value and Deep-Discount Brands (Versus Premium Brands), by Demographic Characteristics and Tobacco Use Patterns.

|  | Value vs. Premium |  | Deep-Discount vs. Premium |  |
| --- | --- | --- | --- | --- |
|  | RPR | (95% CI) | RPR | (95% CI) |
| **Sex** |  |  |  |  |
| Male (*ref*) | 1 |  | 1 |  |
| Female | **1.55\*\*** | **(1.14-2.10)** | 1.12 | (0.77-1.62) |
| **Age** |  |  |  |  |
| 18-25 (*ref*) | 1 |  | 1 |  |
| 26-34 | 1.08 | (0.72-1.61) | 0.87 | (0.48-1.58) |
| 35-49 | **1.62\*** | **(1.03-2.55)** | 1.42 | (0.84-2.40) |
| 50-64 | **2.20\*\*** | **(1.26-3.84)** | **2.15\*\*** | **(1.25-3.70)** |
| 65+ | **4.16\*\*\*** | **(2.13-8.12)** | **5.52\*\*\*** | **(2.79-10.92)** |
| **Race/Ethnicity** |  |  |  |  |
| NH White (*ref*) | 1 |  | 1 |  |
| NH Black | **0.34\*\*\*** | **(0.22-0.54)** | **0.12\*\*\*** | **(0.04-0.34)** |
| Hispanic | 0.69 | (0.39-1.23) | 0.51 | (0.25-1.06) |
| NH Other | 0.72 | (0.40-1.32) | 0.57 | (0.28-1.14) |
| **Income** |  |  |  |  |
| <\$20,000 | **2.22\*\*** | **(1.29-3.83)** | **2.69\*\*** | **(1.50-4.80)** |
| \$20,000 - \$49,999 | **1.85\*** | **(1.13-3.01)** | **2.27\*\*** | **(1.32-3.91)** |
| \$50,000 - \$74,999 | 1.09 | (0.63-1.88) | 1.51 | (0.82-2.78) |
| ≥\$75,000 (*ref*) | 1 | | 1 | |
| **Receives Govt Assistance** |  |  |  |  |
| No (*ref*) | 1 |  | 1 |  |
| Yes | 1.11 | (0.80-1.54) | **1.72\*\*** | **(1.18-2.51)** |
| **Days Smoked (Past 30 Days)** |  |  |  |  |
| Non-Daily (<30 Days) (*ref*) | 1 |  | 1 |  |
| Daily (All 30 Days) | 1.17 | (0.77-1.76) | **1.78\*** | **(1.02-3.08)** |
| **Mean Cigarettes/Day** |  |  |  |  |
| <10 (*ref*) | 1 |  | 1 |  |
| 10 to 19 | **1.76\*** | **(1.06-2.94)** | 1.75 | (0.97-3.13) |
| 20 (1 pack) | 1.72 | (0.92-3.21) | 1.53 | (0.77-3.03) |
| >20 (>1 pack) | 1.90 | (0.85-4.26) | **2.34\*** | **(1.22-4.50)** |
| **Nicotine Dependent** |  |  |  |  |
| No (*ref*) | 1 |  | 1 |  |
| Yes | 1.21 | (0.93-1.58) | **2.26\*\*\*** | **(1.58-3.25)** |

**Bolded RPRs** indicate statistical significance (\*p<0.05, \*\*p<0.01, \*\*\*p<0.001)
<sup>†</sup>Adjusted for sex, race/ethnicity, income, receiving government assistance, daily smoking (past 30 days), mean number of cigarettes smoked per day (past 30 days, categorical), and nicotine dependence.

## DISCUSSION

Using nationally representative data from 2021, this study found that receiving government assistance, heavier smoking, and meeting criteria for nicotine dependence were associated with increased likelihood of using deep-discount—but not value— cigarettes (versus premium). More research is needed to better understand how deep-discount cigarettes may impact tobacco use behavior (including smoking cessation), particularly among older and lower-income adults, who are more likely to smoke cheaper cigarettes.

## Data Availability

All data produced in the present study are available upon reasonable request to the authors.

https://www.samhsa.gov/data/data-we-collect/nsduh-national-survey-drug-use-and-health/datafiles/2021

