## Appendix A for "Sociodemographic characteristics and tobacco use patterns associated with using premium, value, and deep-discount cigarettes among US adults who smoke: A cross-sectional analysis of data from the National Survey on Drug Use and Health, 2021"

**APPENDIX A: List of Brands Categorized as Premium, Value, and Deep-Discount***

| **Premium** | Benson & Hedges, Camel, Capri, Kool, Marlboro, Merit, More, Nat Sherman, Natural American Spirit, Newport, Parliament, Salem, Saratoga, Virginia Slims, Winston |
| --- | --- |
| **Value** | Basic, Best Value, Carlton, Doral, GPC, L & M, L. Ducat, Lucky Strike, Maverick, Misty, Monarch, Old Gold, Pall Mall, Players, True, USA Gold, Wave |
| **Deep-Discount** | 1^st^ Class, 305’s, Berley, Carnival, Cheyenne, Crowns, Decade, DTC, Eagle, Edgefield, Liggett Select, Montclair, Montego, Native, Pyramid, Seneca, Signal, Sonoma, This, Timeless Time, Traffic, USA, Wild Horse |

***** ”Major tobacco companies” refer to Altria (maker of Marlboro in the US and leading cigarette company in the US) and “Big Four” tobacco companies that sell cigarettes in the US (i.e., British American Tobacco, Imperial Brands, and Japan Tobacco). While Philip Morris International is also a “Big Four” company, it does not currently sell cigarettes in the US.
